# Auditory–Somatosensory Influences on Vocal Motor Control in Primary Muscle Tension Dysphonia

**DOI:** 10.1101/2025.10.23.25338678

**Authors:** Adrianna C. Shembel, Elizabeth D. Young, Julianna Smeltzer, Roozbeh Behroozmand

## Abstract

**Purpose:** Auditory and somatosensory systems jointly support vocal motor control, yet their relative and combined contributions in primary muscle tension dysphonia (pMTD) remain poorly understood. This study examined how auditory and somatosensory feedback influence vocal motor control and adaptation in individuals with and without pMTD.

**Method:** Fifty-one participants (pMTD: n = 10; controls: n = 41) completed a 200-trial altered auditory feedback (AAF) paradigm involving sustained vowel productions with fundamental frequency (*f*_₀_) downward shifts at -100 cents. The task was repeated with nebulized lidocaine to transiently attenuate laryngeal somatosensory input. Data were analyzed to extract the magnitude and direction of vocal adjustments across baseline, ramp, hold, and washout phases.

**Results:** Vocally healthy controls demonstrated robust adaptive vocal response to *f*_₀_ feedback alterations followed by an adaptive after-effect during the washout phase, particularly when somatosensation was intact. In contrast, when somatosensation was intact, individuals with pMTD displayed exaggerated overshooting vocal responses in *f*_₀_, lacked typical after-effect adaptation, and showed greater variability overall. Notably, somatosensory disruption stabilized adaptation in pMTD with nebulized lidocaine.

**Conclusion:** These findings demonstrate that pMTD is characterized by maladaptive reliance on somatosensory input that interferes with normal auditory-motor integration. While vocally healthy speakers use somatosensory cues to stabilize vocal control, in pMTD these cues may act as a source of instability. The results underscore the need for therapeutic approaches targeting sensory-motor integration, including individualized interventions that recalibrate the balance between auditory and somatosensory feedback. This study reframes pMTD as a multisensory integration disorder and opens new directions for mechanism-driven voice therapy.

## INTRODUCTION

Auditory adaptation protocols, such as the altered auditory feedback (AAF) protocol, are widely used to study the role of the auditory sensory system in vocal motor control and sensorimotor integration in both healthy and disordered populations (Behroozmand & Sangtian, 2018; Li et al., 2013; Weerathunge et al., 2022). These protocols generally involve real-time digital manipulation of a person’s voice, in which features such as fundamental frequency (*f_0_*) or formant frequencies are artificially shifted upward or downward and played back through headphones while the person vocalizes. In response to the altered feedback that is heard, individuals adjust their vocalizations in subsequent trials, often responding in the opposite direction of the pitch shift. This process is known as auditory-motor adaptation. In vocally healthy individuals, once the altered feedback is removed and the individual once again hears their original, unmodified vocalizations, a period of continued auditory-motor adaptation is common. These vocalizations remain temporarily altered before returning to baseline, which reflects the vocal system’s ability to adapt and integrate prior auditory feedback to compensate for changes in the environment (Kim et al., 2025).

Interestingly, previous studies have reported that approximately one-third of patients with primary muscle tension dysphonia (pMTD) and benign vocal fold lesions (*e.g.,* vocal nodules) either follow the direction of a pitch shift in AAF paradigms (*i.e.,* they change their vocal output in the same direction as the pitch shift), or exhibit larger-than-expected adaptive responses to auditory feedback in the opposing direction (Abur et al., 2021). These findings suggest variability in vocal pitch adaptation in voice disordered populations and potentially disordered vocal motor control.

However, the auditory system is not the only sensory system involved in vocal motor control and it does not function in isolation. Instead, the auditory system works in tandem with the somatosensory system—an intrinsic monitoring-receptor system capable of detecting the position, movement, and tension of the vocal tract, vocal folds, and other laryngeal muscles. (Bradley, 2000; Foote & Thibeault, 2021; Hammer & Krueger, 2013; Hernández-Morato et al., 2023; Hyodo & Katto, 2006; Leonard & Ringel, 1979). Several studies highlight the critical role of somatosensory feedback in vocal motor control. One study found that anesthesia to the larynx increased vocal instability in vocally healthy individuals, which was reflected in longer and slower transitions between tones during frequency-modulated tone matching tasks, despite no impact on pitch accuracy (Leonard & Ringel, 1979). This study attributed these vocal motor control patterns to laryngeal mucosal receptors and subglottic pressure feedback required for rapid vocal fold adjustments. A second study reported increased acoustic jitter during high-frequency phonation under topical anesthesia in healthy controls and implicated tactile feedback from the laryngeal mucosa to maintain phonatory stability, especially with increased physiological vocal demands (*e.g.,* increased pitch range) (Sorensen et al., 1980). A third study further demonstrated that laryngeal anesthesia disrupted frequency regulation (Tanabe et al., 1975). A fourth study confirmed that laryngeal anesthesia disrupts subglottic pressure regulation and airflow rate stability, linking reduced somatosensory input to impaired fine motor control (Sundberg et al., 1995). Across all these studies, the absence of sensory feedback under laryngeal anesthesia impaired fine motor control, especially when vocal demands were increased, amplifying variability in acoustic vocal output.

These somatosensory vocal motor control findings could also be relevant to clinical populations with functional voice disorders, like pMTD. pMTD is characterized by unstable vocal control, compensatory hyperfunction, and effortful phonation in the absence of structural pathology (Hillman et al., 2020; Roy, 2008; Shembel et al., 2024). Patients with pMTD have been shown to exhibit increased variability with acoustic voicing onset measures (McKenna et al., 2020) and have more variable vocal motor control patterns in the perilaryngeal (Hogue et al., 2023) and laryngeal systems (Crocker et al., 2024). These motor patterns and issues with sensorimotor integration may be due to underlying impairments in fine motor control from disordered somatosensory feedback. Several recent studies have suggested the key role the somatosensory system plays in pMTD. One study found increased somatosensory perceptions to laryngeal tactile stimulation (Shembel et al., 2024), while another found somatosensory awareness differences in patients with pMTD compared to vocally healthy controls (Smeltzer et al., 2023). Multiple studies have observed that patients with pMTD report increased somatosensory voice symptoms even when their vocal motor control patterns are relatively similar between groups with and without pMTD (Hogue et al., 2023; Moore & Shembel, 2025; Morrison et al., 2024; Shembel, Morrison, Fetzer, et al., 2023). However, the role of the somatosensory system in vocal motor control and its relationship to the auditory system requires elucidation in general, and more specifically to patients with pMTD.

Understanding the underlying auditory and somatosensory mechanisms is essential for tailoring precise, individualized voice therapy. Depending on how these sensory systems support optimal vocal motor control in patients with pMTD, voice therapy may need to focus more on the sound (auditory) or the feel (somatosensory) of the voice, with emphasis on the system that most strongly influences the voice disorder. As such, the overarching goal of this study was to study the effects of somatosensory feedback and disruption on auditory-vocal motor control and adaptation in participants with and without pMTD. Because prior studies have often grouped together individuals with varying types of hyperfunctional voice disorders (e.g., pMTD and benign vocal fold lesions)—conditions that may differ in their underlying pathophysiological mechanisms—the present study aimed to more precisely characterize auditory-vocal motor control in patients with pMTD. As such, the first objective of the study was to examine how individuals with and without pMTD (specifically) adapt their vocal pitch in response to a well-established AAF protocol, focusing on both the magnitude and direction of pitch-shift responses.

Building on this, the second objective was to examine how disrupting somatosensory feedback—via nebulized lidocaine applied to the larynx and upper airways—affects vocal motor control and adaptation to pitch-shifted stimuli, and whether this effect differs between groups with and without pMTD. The third objective was to evaluate the interaction between auditory and somatosensory feedback systems in shaping vocal motor control, and to explore whether differences in multisensory integration contribute to the variability in vocal adjustments observed in individuals with pMTD.

## METHODS

### Participants

The study was approved by the University of Texas (UT) Southwestern Medical Center Institutional Review Board (STU-2020-0720) and all participants gave informed consent. The study involved 51 subjects (10 patients with pMTD, 41 vocally healthy controls). Participants with pMTD were identified and recruited from the UT Southwestern Center for Voice Care, while the vocally healthy controls were recruited from the greater Dallas-Fort Worth region. To be eligible for the pMTD group, participants needed to have a formal diagnosis of pMTD from a board-certified laryngologist and confirmed by a voice-specialized speech-language pathologist. Participants with pMTD also needed to report significant vocal impact on the Voice Handicap Index-10 (Rosen et al., 2004) (VHI-10 mean = 15.56 ±7.09), vocal fatigue based on Part 1 of the Vocal Fatigue Index (Nanjundeswaran et al., 2015) (VFI-1 mean = 25.00 ±8.99), and vocal tract discomfort based on the Vocal Tract Discomfort Scale (Lopes et al., 2015; VTDS mean = 28.86 ±16.12). These inclusion criteria have previously been used across multiple other studies from our group (Hogue et al., 2023; Morrison et al., 2024; Shembel et al., 2024; Shembel, Morrison, Fetzer, et al., 2023; Shembel, Morrison, McDowell, et al., 2023) and involve the most salient voice symptoms in patients with pMTD (Moore & Shembel, 2025). Participants could also exhibit presence of mediolateral or anteroposterior supraglottic compression, as well as aberrant vocal quality/acoustic vocal output. However, these laryngoscopic, auditory-perceptual, and acoustic features were not considered in the inclusion criteria because not everyone with pMTD has these clinical presentations (Shembel et al., 2021; Shembel, Morrison, McDowell, et al., 2023).

### Data Collection

All participants underwent the AAF protocol twice. The first protocol block was always without lidocaine (Block 1-Sensation Intact). Before beginning the official study, 11 participants in the vocally healthy group were first randomly selected to complete the protocol without lidocaine in both blocks (Block 1-Sensation Intact / Block 2-Sensation Intact; Figure 1). This study design was selected to verify that presenting the AAF protocol twice did not introduce an order effect, and to ensure that any differences between block conditions were attributable to lidocaine rather than presentation order. Because the effects of lidocaine can last up to two hours (Beecham et al., 2025), randomizing the order of the two conditions to do a true cross-over study design would have required two separate testing days, which would have unnecessarily increased participant attrition. Figure 1 displays the adaptation responses of participants after completing two testing blocks with sensation intact. A visual analysis demonstrates that adaptation followed the expected pattern for vocally healthy controls in both blocks—that is, vocally healthy controls first changed their vocal *f_0_* output in the opposite direction and then adapted to the pitch shifts during the hold and washout periods. To further confirm the utility of the study design, a Fisher’s exact test comparing the number of outliers in each testing blocks were conducted based on prior literature (Kapsner-Smith et al., 2024). The results were non-significant (*p* = 0.670), implicating that participants who were outliers in the first block remained outliers in the second block and that order effect did not play a role in their responses. Given these findings on the lack of order effect of conducting the AAF protocol twice, the study proceeded with all 10 participants with pMTD and 30 additional controls, each completing two blocks: Block 1 (Sensation Intact) followed by Block 2 (Lidocaine).

**Figure 1:**
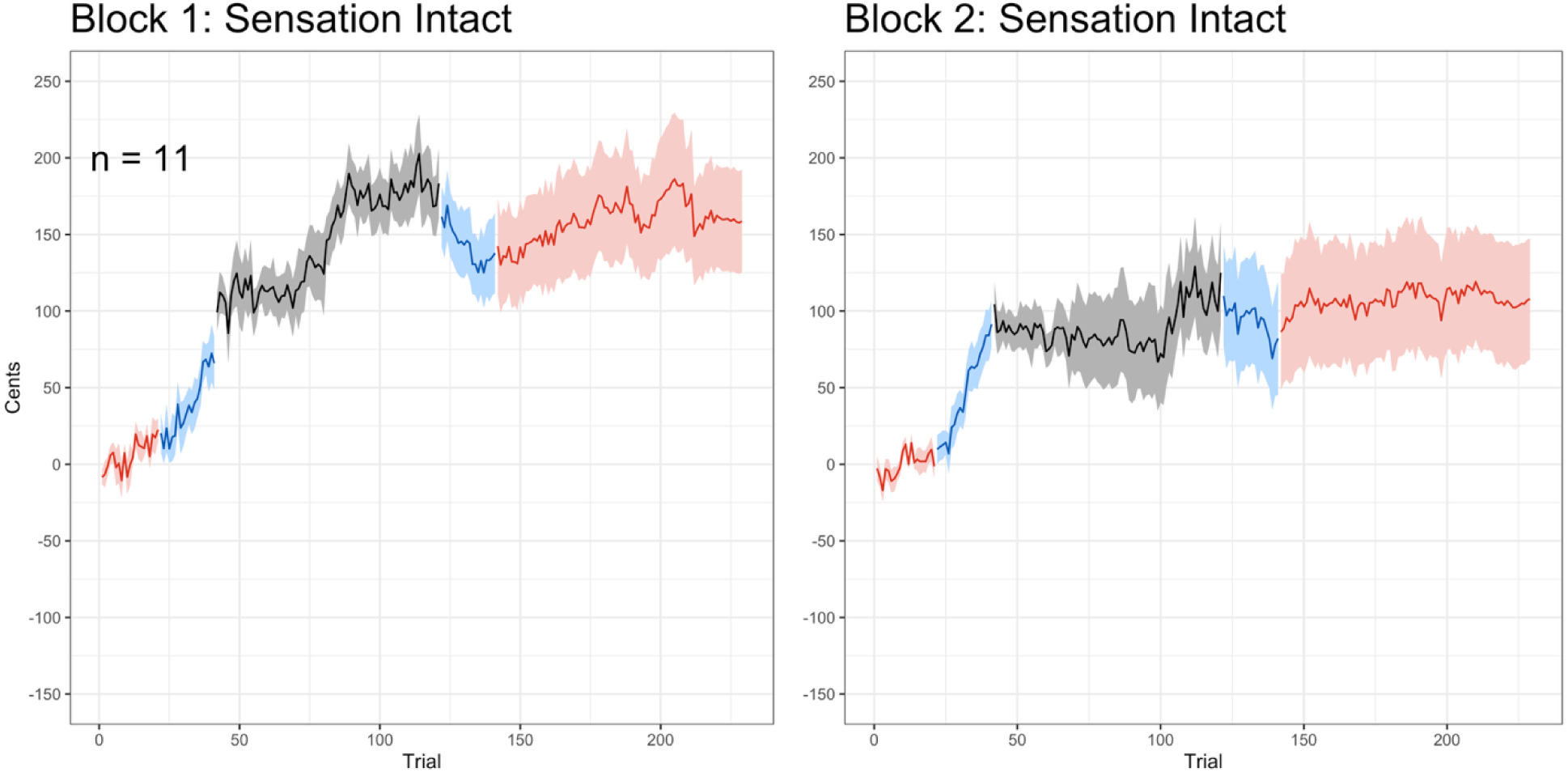
Verification of the absence of an order effect for the altered auditory feedback stimulus presented twice

#### Altered Auditory Feedback (AAF)

The AAF protocol used for this study is an established vocal motor control paradigm to study adaptations to real-time *f_0_* shift perturbations in auditory feedback (Behroozmand et al., 2012; Behroozmand & Sangtian, 2018; Li et al., 2013; Weerathunge et al., 2022). Participants were asked to produce the speech vocal sound /i/ (as in the word “neat”), each held for 3-5 seconds in each trial (Fig 2A), while auditory feedback involving downard *f_0_* shifts at -100 cents magnitude or no *f_0_* shifts were presented to participants through headphones (Fig 2B). Participants were first fitted with a head-mounted AKG condenser headset (model HSC 271), connected to a Motu Ultralite-MK3 Hybrid sound card and amplifier that was used to record voice samples at a 44.1 kHz sampling rate on a laboratory computer. The *f_0_* shifts were conducted using the H90 Eventide Harmonizer. The first set of 20 trials involved no shifts in the auditory feedback (Pre-shift [Baseline] in Fig 2C). This was immediately followed by a second set of 20 trials involving gradual downward shifts in *f_0_* by 5 cents (*f_0_* Ramp Down in Fig 2C). The third set included 80 trials with *f_0_* shifts maintained at -100 cents (Maintenance [Hold] in Fig 2C). The following fourth set of 20 trials included gradual *f_0_* shifts upwards by 5 cents (*f_0_* Ramp Up in Fig 2C). The fifth set included 60+ trial presentations of unaltered (no *f_0_*shift) auditory feedback (Post-Shift [After-Effect] in Fig. 2C).

**Figure 2:**
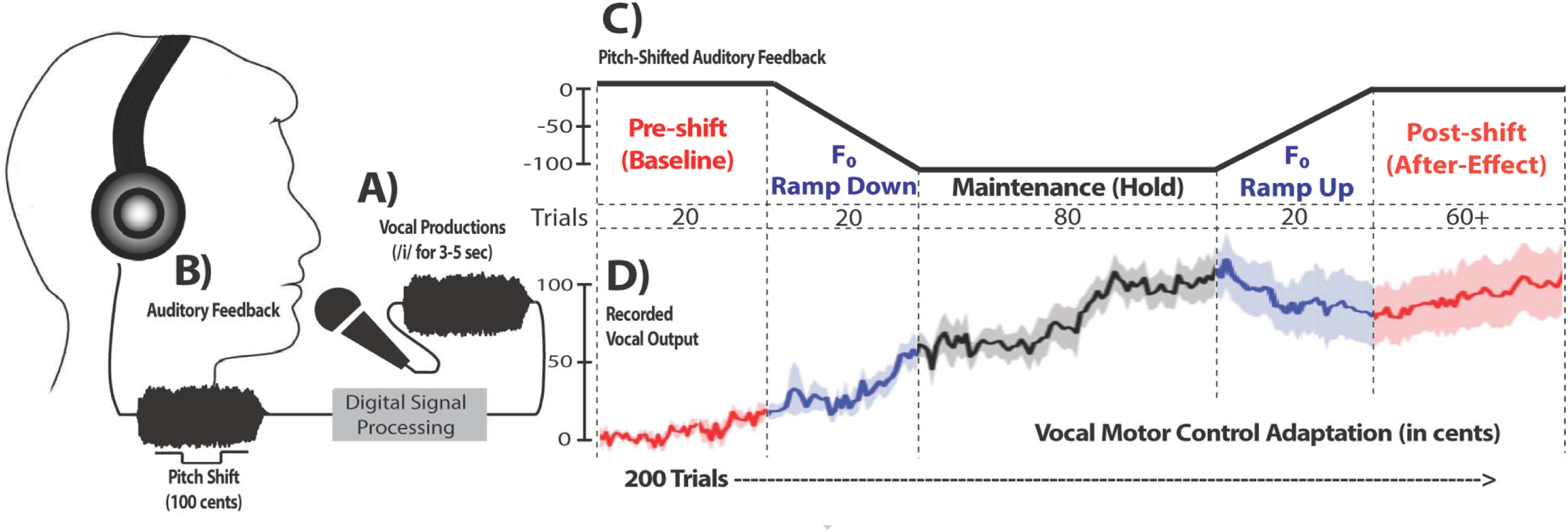
Altered Auditory Feedback Paradigm and Setup Schematic

#### Nebulized Lidocaine

After the first block was completed, 3 cc of 4% nebulized lidocaine was administered via a jet nebulizer and mouthpiece for 3-5 minutes. A compression nebulizer (Henry Schein, model 112-7063) was used to aerosolize the solution. Participants were asked to take deep, slow breaths through their mouth to allow effective deposition of the aerosolized lidocaine into the larynx and upper airways. Lack of sensory input to the larynx was confirmed via a flexible laryngoscope in five healthy controls to ensure its utility. This method has also been previously used in the literature to study vocal motor control (Yang & Chen, 2005) and is a standard of care numbing protocol for in-office laryngeal surgical procedures. Once participants were nebulized, they were asked to complete another round of the AAF protocol (Block 2-Lidocaine) in the same fashion as Block 1-Sensation Intact. Each block took 15-20 minutes to complete, which is well within the lidocaine activation range.

### Data Processing

Measures of vocal motor adaptation responses to auditory feedback *f_0_*-shift stimuli were extracted following standardized procedures described in previous studies (Behroozmand et al., 2012; Behroozmand & Sangtian, 2018). The single-trial pitch frequency contours were first extracted from the recorded voice signals in Praat (Boersma, 2001) and a customized MATLAB code. Voice frequencies during vocalization trials for each phase (baseline, ramp down, hold, ramp up, and after-effect were measured separately. For each trial, vocal frequency changes were converted from Hertz to Cents using the following formula: 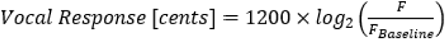, where F is the frequency of a single vocal production trial during each phase relative to the voice frequency averaged across all pre-shifted baseline trials (F_Baseline_). The extracted vocal responses in Cents for each participant were averaged and compared across all participants during each phase.

### Data Analysis

Linear-mixed effects models were used to examine the effects of group (pMTD vs. control) and somatosensory feedback condition (Block 1-Sensation Intact, Block 2-Lidocaine), as well as their interaction, on the adaptive vocal motor responses to pitch-shift stimuli. All statistical analyses were completed using R (version 4.3.3). The linear mixed effects models were conducted using the *lmerTest* package (version 3.1-3). Significant interactions were stratified using the *emmeans* package (version 1.10.0). The overall effect of group, condition, and the group x condition interaction was examined (*i.e.,* overall adaptation across all phases), as well as the effect of group, condition, and the group x condition interaction for the hold phase (*i.e.,* the degree of adaptation with maximal altered auditory feedback) and the after-effect phase (*i.e.,* the degree of adaptation following removal of altered auditory feedback). Random effect structures were specified based on a bottom-up approach, where theoretically-sound random effects were added one at a time until the model did not converge or the addition of a random effect did not significantly improve model fit as assessed by a -2 log-likelihood test using the *anova* function from the base *stats* package. In all models, only a random intercept for participants was included; attempts to include random slopes for participant group were not supported by the data. The percent of variance explained compared to a null model (*i.e.,* a model containing only the random intercept of participant) was calculated for each model as a measure of effect size, using the following formula: (residual_null model–_ residual_current model_)/residual_null_ model (Peugh, 2010).

In addition, chi-squared tests of homogeneity were used to examine associations between groups and individual participant responses. Chi-square tests were conducted using the base s*tats* package in R. Each participant’s overall response was categorized as either (1) following the downwards pitch shift below the 90^th^ percentile z-cutoff of the control group in each condition, (2) increased magnitude of opposing upward shifts defined by above 90^th^ percentile z-score cutoff of the control group in each condition, or (3) typical (within the range of the control group in each condition), as has been done in previous work (Abur et al., 2021; Kapsner-Smith et al., 2024).

## RESULTS

The response of each participant group by condition is displayed in Figure 3. There was a significant effect of condition (β = -15.87, t(18600) = -11.29, *p <* .001) where the mean adaptation in the sensation intact (no lidocaine) condition was 42.2 cents compared to 58.0 cents in the lidocaine condition. While the main effect of group was not significant, (β =-1.12, t(40)= -0.04, *p* = .970) a significant condition x group interaction was found (β = -32.39, t(186000) = 10.00, *p* < .001). The estimated means were examined using Sidak corrections for multiple comparisons. Only the difference between the pMTD group for the sensation intact vs lidocaine condition was significant (approximately 40 cents higher in the lidocaine condition; *p <* .001) as well as the difference for the control group for the sensation intact vs lidocaine condition (approximately 7 cents higher in the lidocaine condition; *p <* .001). While other contrasts visually appeared quite different, the high variability within each condition likely contributed to their lack of statistical significance (see Figure 4).

**Figure 3:**
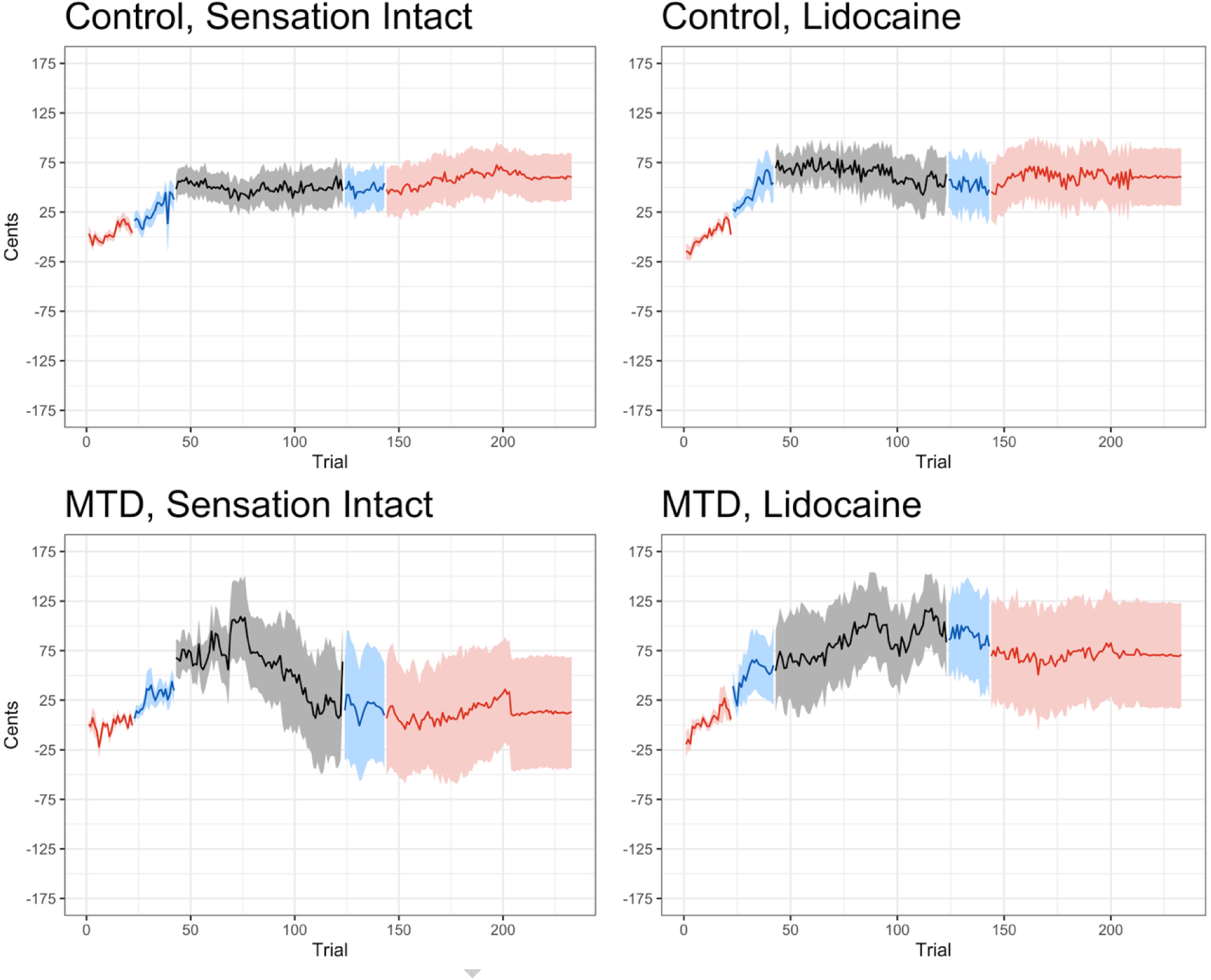
Responses to altered auditory feedback paradigm by participant group (control [top panels], primary Muscle Tension Dysphonia [bottom panels]) and condition (sensation intact [left panels], lidocaine [right panels]). In all figures, baseline and after-effect phases are represented in red, ramp up and ramp down phases are represented in blue, and hold phases are represented in black. The solid line in each figure represents the group average, and the shaded area around each solid line represents the standard error.

**Figure 4:**
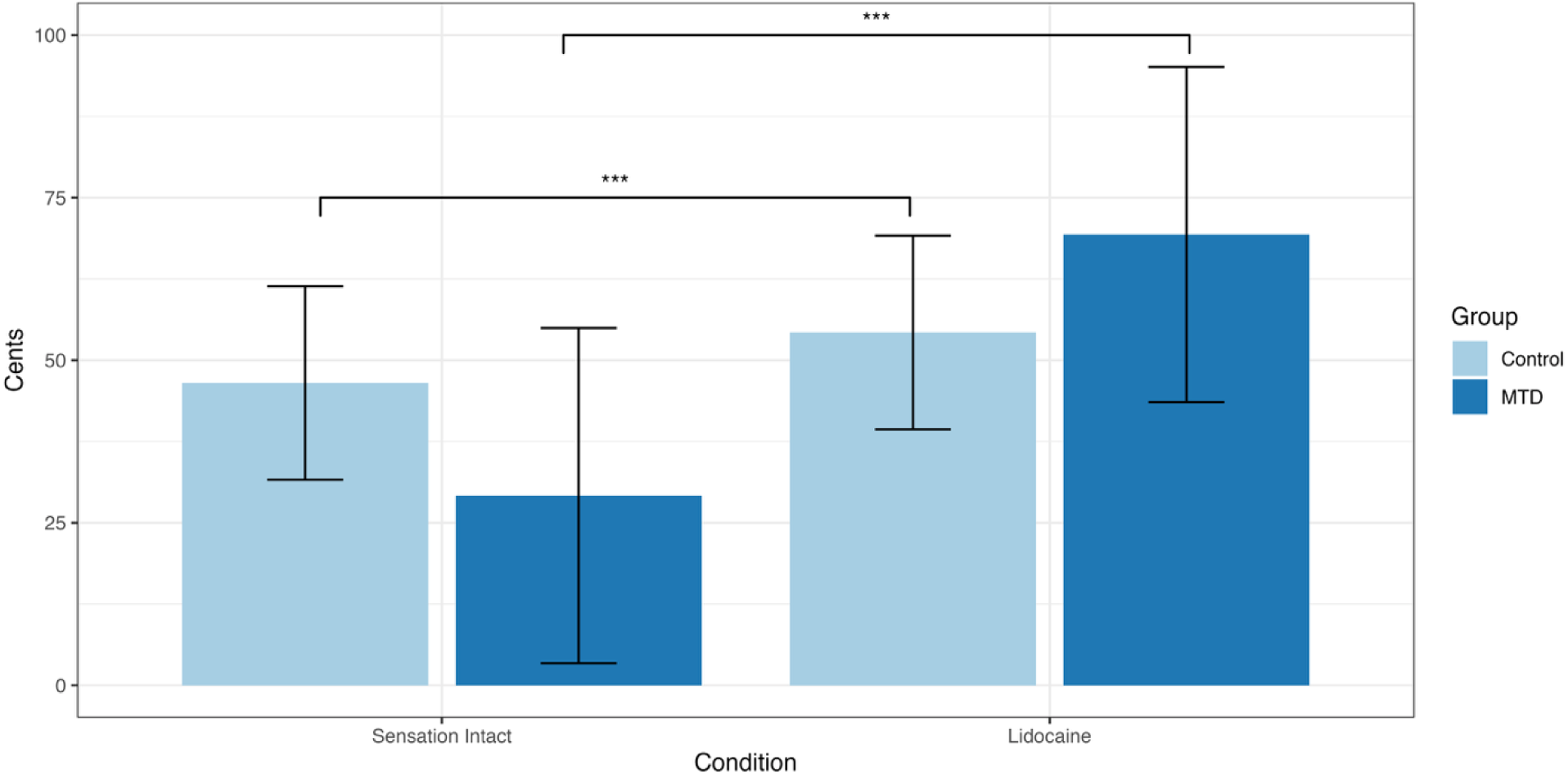
Interaction effect between condition (sensation intact vs nebulized lidocaine) and group (vocally healthy control vs primary MTD). Error bars represent standard error.

Additionally, there was a significant interaction between group and condition in the adaptation phase, β = -10.82, t(6440) = -2.34, *p =* 0.019. Similar to the trend found across all phases, stratification of this interaction indicated that there was a significant difference between the pMTD group for the sensation intact vs lidocaine conditions as well as the control group for the sensation intact vs lidocaine conditions. There was also a significant group x condition interaction in the after-effect phase, β = -56.38, t(7160) = -11.49, *p* < .001; within this interaction, only the difference between the pMTD group for the sensation intact vs lidocaine conditions reached significance (approximately 58 cents higher in the lidocaine condition; *p <* .001). The percent of variance explained by each of the models described above is presented in Table 1. Means and standard deviations across group and condition are represented in Table 2.

**Table 1.** Proportion of variance explained for linear mixed-effects models.

| Model | Model AIC | Model Residual (SD) | $\Delta$ Residual | Variance Explained |
| --- | --- | --- | --- | --- |
| Null Model – All Phases | 223414.8 | 9275 (96.31) | - | - |
| Fixed effect of Condition | 223289.8 | 9212 (95.98) | 63 | 0.68% |
| Fixed effect of Group | 223416.8 | 9275 (95.98) | 0 | 0% |
| Group x Condition Interaction | 223194 | 9163 (95.72) | 112 | 1.21% |
| Null Model – Hold Phase Only | 75584.7 | 6597 (91.22) | - | - |
| Group x Condition Interaction | 75507.7 | 6515 (80.71) | 82 | 1.24% |
| Null Model – After-Effect Phase Only | 85670.2 | 8333 (91.28) | - | - |
| Group x Condition Interaction | 85491.3 | 8120 (90.11) | 213 | 2.56% |

Notes: Null models contained only a random intercept of participant. Proportion of variance explained was calculated separately for each set of models (i.e., All phases, Hold phase only, After-effect phase only).

**Table 2.** Mean and Standard Deviation Responses in Cents by Condition, Group, and testing Phase.

| Condition | Group | Phase | Mean (cents) | SD (cents) |
| --- | --- | --- | --- | --- |
| Sensation Intact | Control | Baseline | 4 | 31 |
| Sensation Intact | Control | Ramp Up | 48 | 112 |
| Sensation Intact | Control | Hold | 49 | 94 |
| Sensation Intact | Control | Ramp Down | 26 | 64 |
| Sensation Intact | Control | After-Effect | 59 | 130 |
| Lidocaine | Control | Baseline | 1 | 31 |
| Lidocaine | Control | Ramp Up | 52 | 156 |
| Lidocaine | Control | Ramp Down | 45 | 79 |
| Lidocaine | Control | After-Effect | 60 | 159 |
| Lidocaine | Control | Hold | 65 | 121 |
| Sensation Intact | pMTD | Baseline | 0 | 21 |
| Sensation Intact | pMTD | Ramp Up | 18 | 175 |
| Sensation Intact | pMTD | Hold | 59 | 126 |
| Sensation Intact | pMTD | Ramp Down | 28 | 36 |
| Sensation Intact | pMTD | After-Effect | 12 | 171 |
| Lidocaine | pMTD | Baseline | 4 | 30 |
| Lidocaine | pMTD | Ramp Up | 90 | 138 |
| Lidocaine | pMTD | Hold | 85 | 125 |
| Lidocaine | pMTD | Ramp Down | 51 | 82 |
| Lidocaine | pMTD | After-Effect | 70 | 160 |

The number of participants within each group that were classified as typical and atypical responders using a 90^th^ percentile cut-off are displayed in Table 3. There was no difference in the proportion of atypical responders between control and pMTD groups in either the Sensation Intact or Lidocaine conditions (see Table 3).

**Table 3.**
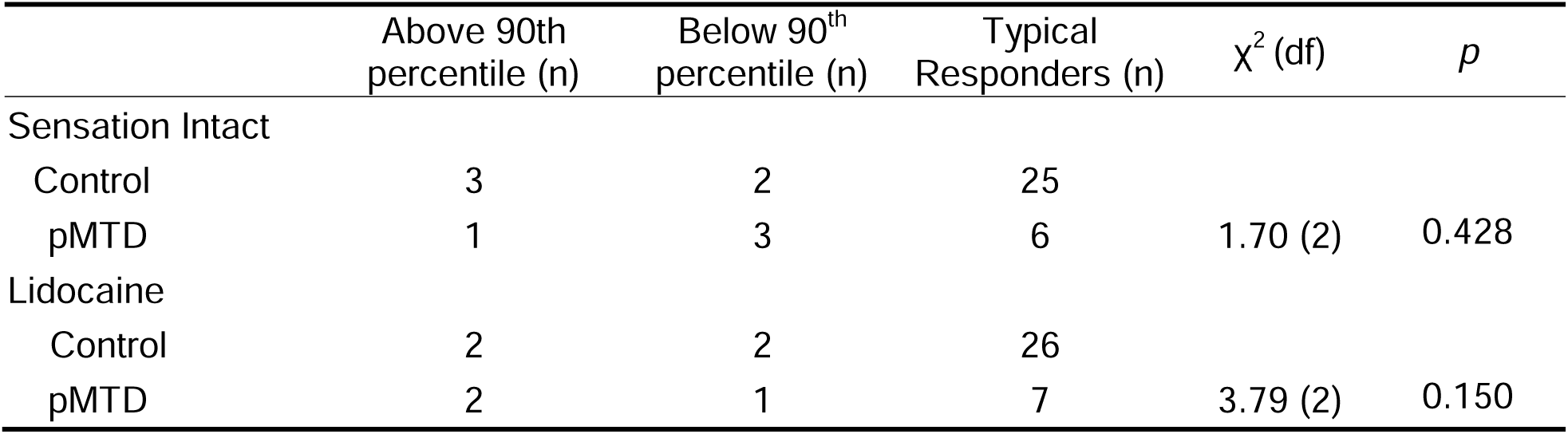
Number of Typical and Atypical Responders by Condition and Group.

## DISCUSSION

The present study investigated how auditory and somatosensory systems contribute to vocal motor control and adaptation in individuals with and without pMTD. Using an AAF paradigm with and without laryngeal anesthesia, the three study objectives were to determine (1) whether individuals with pMTD adapt differently to pitch-shifted feedback compared to healthy controls, (2) whether disrupting somatosensory input modulates auditory-driven adaptation, and (3) how interactions between auditory and somatosensory systems influence vocal motor control across groups.

### Auditory-Motor Adaptation in Vocally Healthy Controls and pMTD

Consistent with prior studies, vocally healthy controls adapted to downward *f_0_* shifts in the shift-down phase by raising their vocal pitch in the hold and after-effect phases, in parallel with previous studies (Behroozmand & Sangtian, 2018; Kim et al., 2025; Li et al., 2013). This robust pattern of compensation and adaptation underscores the stability and flexibility of auditory-motor integration in vocally healthy systems. In contrast, individuals with pMTD displayed markedly different patterns of responses. They tended to exhibit overshooting vocal responses during the ramp down phase and failed to exhibit the typical adaptation in the after-effect phase once pitch shifts were removed. This aberrant vocal response pattern and lack of adaptation suggest alterations in auditory input and vocal motor output in these individuals. These auditory-motor patterns may reflect either disrupted sensory integration or impaired sensorimotor control calibration. Interestingly, these individuals also had more varied responses, demonstrated by the greater variability (*i.e.,* higher standard deviations and standard error; see Figures 3 and 4) across all phases, which could further explain the unstable vocal motor control systems previously observed in patients with pMTD (Crocker et al., 2024; Hogue et al., 2023; McKenna et al., 2020). These findings align with previous reports of atypical and varied pitch-shift responses in individuals with hyperfunctional voice disorders (pMTD, benign vocal fold lesions) (Abur et al., 2021), and extends this work by isolating the specific contributions of auditory feedback on vocal motor control in individuals with pMTD rather than collapsing across multiple hyperfunctional voice disorders.

### Effects of Somatosensory Disruption in Vocally Healthy Controls and pMTD

A novel aspect of this study was the examination of somatosensory disruption via nebulized lidocaine in individuals with and without pMTD. In vocally healthy controls, reduced laryngeal sensation produced an increase in compensatory pitch, consistent with prior work showing that somatosensory deprivation can exaggerate corrective responses (Larson et al., 2008). With intact somatosensation, corrective responses to altered auditory feedback are present but relatively modest because the brain can cross-check auditory errors against stable somatosensory input. However, with somatosensory disruption, corrective responses in the opposing direction become larger and exaggerated because the nervous system relies more heavily on auditory error signals in the absence of reliable information and stable references in the tactile and proprioceptive system. In other words, somatosensory deprivation removes an essential sensory anchor, resulting in an auditory mismatch that feels more uncertain, prompting exaggerated corrections. These findings reinforce the interdependent roles of auditory and somatosensory systems in stabilizing vocal motor control.

Interestingly, individuals with pMTD overshot in the opposing direction in both condition blocks (sensory intact, lidocaine) compared to vocally healthy controls. Individuals with pMTD also had more stable adaptation in the after-effect phase with lidocaine relative to their sensation intact block, and these adaptive patterns were more like the vocally healthy controls. These findings suggest that in individuals with pMTD, somatosensory input may contribute to maladaptive vocal motor patterns, such that temporarily reducing its influence enables more normalized auditory-driven adaptation. These findings align with growing evidence that heightened or dysregulated somatosensory feedback and processing is a feature of pMTD (Moore & Shembel, 2025; Shembel et al., 2024; Smeltzer et al., 2023).

Although significant group differences were observed in mean vocal motor control and adaptation, no significant differences emerged in the *proportion* of atypical responders between individuals with pMTD and healthy controls across either condition. A recent analysis of auditory perturbation paradigms in both healthy individuals and patients with Parkinson’s disease found no evidence for a distinct subgroup of “followers”—that is, atypical responders who follow the direction of the pitch perturbation rather than compensating in the opposite direction (Miller et al., 2023). The authors of that review proposed that following responses likely represent the tails of a normal distribution rather than a qualitatively unique response pattern. The non-significant chi-square tests in the current study align with this interpretation. However, it is noteworthy that 30–40% of participants with pMTD (depending on condition) were classified as atypical responders—a proportion higher than would be expected under a normal distribution, yet consistent with previous findings in this population (Stepp et al., 2017; Abur et al., 2021, 2023).

### Multisensory Integration and Variability in pMTD

The significant group by condition interaction underscores that auditory and somatosensory systems interact differently in individuals with pMTD compared to vocally healthy controls. Whereas healthy controls rely on somatosensory input to fine-tune vocal motor control responses, individuals with pMTD may experience interference from aberrant or over-weighted somatosensory signals. This sensory imbalance may help explain the overshoot in vocal compensation, greater variability observed in pMTD patients’ compensatory behaviors, and broader difficulties with adaptation and vocal stability. The findings support a model of pMTD as a sensory-motor disorder, rather than one rooted solely in excessive muscular hyperfunction.

### Clinical and Theoretical Implications

These findings have several important implications. First, they suggest that effective intervention for pMTD may require not only muscle unloading but also vocal techniques that involve sensory discrimination and sensorimotor integration. These findings could explain why voice therapies that focus on somatosensory discrimination and integration (*e.g.,* resonant voice therapy) are successful in clinical populations with pMTD. Additional voice therapy techniques could include recalibration of the auditory-motor system (*e.g.,* structured auditory perturbation exercises) or approaches that modulate somatosensory input (*e.g.,* tactile biofeedback, kinesthetic awareness training) to rebalance multisensory control of phonation. Second, these results highlight the importance of individualized approaches for patients with pMTD. For example, some patients may benefit from strategies emphasizing auditory cues (*e.g.,* the sound of the voice), while others may require interventions that reshape maladaptive somatosensory reliance (*e.g.,* the feel of the voice). Specific techniques that target the auditory and somatosensory systems during stimulability testing to determine optimal treatment approaches for individuals with pMTD also require further development. On a theoretical level, this study reinforces the need to conceptualize voice disorders within a multisensory integration framework, consistent with broader models of motor control in speech and movement systems.

### Limitations and Future Directions

Several limitations should be acknowledged. One limitation concerns the modest sample size of the pMTD group, potentially reducing the study’s statistical power and limiting the generalizability of its conclusions. However, effects with and without lidocaine were still observed in this smaller pMTD cohort, suggesting findings are relevant to this clinical population. A related limitation is the small proportion of variance explained by the statistical models within the current study (see Table 1). This is likely due to the inherent variability of a pitch adaptation altered auditory feedback task, as well as the substantial variability seen with the pMTD cohort. The use of mixed-effects modeling is an effective method of accounting for individual variability for this paradigm compared to traditional ANOVA models, as it accounts for the covariance between each individual and their own datapoints (Quené & van den Bergh, 2004). A second limitation is that while the nebulized lidocaine protocol provided a controlled model of transient somatosensory disruption, it may not fully capture the chronic sensory processing differences present in individuals with pMTD.

The third potential limitation was order effect with and without lidocaine. The AAF protocol was always administered without lidocaine prior to conducting the protocol with lidocaine. This order was chosen to ensure that the initial block represented a true baseline condition. Lidocaine can have lingering physiological effects on sensory function, which could confound subsequent measures. By always starting with the sensation intact (*i.e.,* no lidocaine) condition in the first block, we ensured that the comparison between baseline and lidocaine-altered states was not biased by carryover effects. A limitation of this fixed order is that it introduces the possibility of order effects—such as learning, adaptation, or fatigue—that could contribute to differences observed between conditions. To address this concern, we conducted a control experiment in which healthy participants completed two consecutive AAF blocks without lidocaine. These additional blocks allowed us to estimate the extent of order-related changes independent of lidocaine, strengthening confidence that the effects observed in the main protocol were not solely attributable to order. Nonetheless, the inability to fully counterbalance condition order remains a methodological limitation. The fourth limitation is that the study only focused on behavioral responses. Future research should integrate neural measures (e.g., EEG, fMRI) to more directly probe the mechanisms of auditory-somatosensory integration as it relates to the vocal motor control system in clinical populations with pMTD. Finally, longitudinal studies are needed to evaluate whether targeted sensory-motor interventions can normalize vocal motor control adaptation responses and reduce clinical voice symptoms.

## CONCLUSION

This study provides critical evidence that individuals with pMTD differ fundamentally from vocally healthy controls in the way auditory and somatosensory systems interact to regulate vocal motor control. In vocally healthy controls, intact somatosensory input serves as a stabilizing reference point that tempers corrective responses to auditory perturbations, thereby ensuring vocal stability. By contrast, individuals with pMTD demonstrate maladaptive reliance on somatosensory feedback, such that attenuation of these inputs via laryngeal anesthesia facilitated more normalized auditory-driven adaptation. These findings underscore the role of disordered or over-weighted somatosensory processing in the pathophysiology of pMTD.

Clinically, these findings highlight the need for therapeutic approaches that extend beyond strategies for muscular unloading and address the sensory-motor foundations of pMTD. Treatment paradigms that incorporate somatosensory discrimination training, tactile biofeedback, or structured auditory perturbation exercises may help recalibrate maladaptive integration patterns and restore more balanced vocal motor control. Importantly, the heterogeneity of responses observed in the pMTD group suggests that therapy should be individualized, with consideration of whether a patient is more reliant on auditory or somatosensory information.

Theoretically, this study advances our understanding of pMTD as a disorder of multisensory integration rather than one solely rooted in hyperfunctional muscular activity. By situating pMTD within broader models of sensory-motor control, this work lays the groundwork for future research examining neural mechanisms of auditory-somatosensory interactions. Future work should extend this line of inquiry with larger sample sizes to fully capture the breadth of pMTD heterogeneity. Longitudinal and neuroimaging studies are also warranted to determine whether recalibrating sensory integration through therapy can yield durable improvements in vocal stability and symptom reduction.

## Data Availability Statement

All data are available by request from the authors.

## Data Availability

All data produced in the present study are available upon reasonable request to the first author

